# Interval colorectal cancer risk by haemoglobin concentration in Australia’s National Bowel Cancer Screening Program

**DOI:** 10.64898/2026.10.26.26366479

**Authors:** Driss Ait Ouakrim, James Dowty, Jennifer McIntosh, Belinda Goodwin, Carlene Wilson, Katelyn Collins, Nick Ormiston-Smith, Pantea Konn, Glenn Austin, Eleonora Feletto, Jie-Bin Lew, Uri Ladabaum, Finlay Macrae, Nancy N. Baxter, Mark Jenkins

## Abstract

**Background:** Interval colorectal cancers (CRCs) are an important measure of screening performance. We assessed interval CRC risk after negative faecal immunochemical testing (FIT), its association with faecal haemoglobin (f-Hb) concentration, and differences by age and sex in Australia s National Bowel Cancer Screening Program (NBCSP).

**Methods:** We conducted a population-based retrospective cohort study linking Queensland screening and cancer registry records for NBCSP participants aged 50-74 years screened in 2018-2022. Eligible episodes had f-Hb below 20 ug/g in both samples. We excluded episodes with CRC diagnosed on or before follow-up start. Follow-up ended at CRC, the next screening episode, death, 24 months, or December 31, 2023. We estimated adjusted cause-specific hazard ratios (HRs), accounting for repeated episodes within participants, and cumulative incidence with death and subsequent screening as competing events.

**Findings:** Among 1,083,149 negative screening episodes from 700,734 participants, 586 interval CRCs occurred over 2,056,405 person-years. The incidence rate was 28.5 per 100,000 person-years, and 24-month cumulative incidence was 57.6 per 100,000 episodes (95% CI 53.1-62.5). Risk increased with age and was higher in males (adjusted HR 1.28, 95% CI 1.08-1.50). The complete-case quantitative cohort included 326,931 episodes and 138 cancers. Compared with maximum f-Hb of 0-5 ug/g, concentrations >15-<20 ug/g were associated with an adjusted HR of 14.93 (9.02-24.70). Corresponding 24-month risks were 35.7 (28.5-44.7) and 504.5 (323.4-787.0) per 100,000 episodes. Higher negative-range f-Hb concentrations were more common in older participants and males. The quantitative cohort showed no clear sex difference after adjustment for f-Hb.

**Interpretation:** CRC risk varies substantially among people with a negative screening result. Quantitative F-Hb levels may help identify who is at higher risk of interval CRC. Further research is needed to confirm these findings and assess the benefits, harms and costs of changing screening intervals.

**Research in context:** *Research in context:* Evidence before this study
We searched PubMed October 2026, for Australian studies and international systematic reviews and meta-analyses, using combinations of “colorectal”, “bowel”, “interval cancer”, “interval cancers”, “interval colorectal”, “negative FIT”, “faecal haemoglobin”, “fecal hemoglobin”, “faecal immunochemical”, and “fecal immunochemical”, combined with “Australia”, “Australian”, “systematic review”, or “meta-analysis”. Additional searches examined international cohort studies published from January 1, 2020, to October 25, 2026. Australian national monitoring and a study in Western Australia showed that interval CRC was uncommon but more frequent among males and older participants. Both analyses were based on the 2018 screening cohort, before biennial screening had been fully implemented for all eligible people aged 50-74 years. Neither examined interval CRC risk across the negative f-Hb range. International reviews documented substantial variation in interval CRC incidence between screening programmes. More recent Danish and Dutch cohort studies showed that higher f-Hb concentrations below the positivity threshold were associated with greater interval CRC risk, including after successive negative screening rounds. A dose-response meta-analysis of 13 studies supported the prognostic value of negative-range f-Hb, but combined evidence on interval cancers, cancers detected at subsequent screening, and adenomas. These outcomes do not provide interchangeable estimates of risk before the next screen. The evidence therefore supported further investigation of quantitative f-Hb, but did not establish concentration-specific interval CRC risks for the NBCSP’s two-sample protocol and 20 µg/g positivity threshold. Added value of this study
This study extends Australian evidence through longitudinal linkage of Queensland screening and cancer registry records from 2018-2022, including repeated screening episodes from more than 700,000 participants. It estimates interval CRC risk overall and by age and sex, and examines how that risk varies within the negative f-Hb range. Follow-up ended at the next screening episode, allowing interval cancers to be distinguished from cancers detected at later screening. The quantitative analysis showed a substantial concentration gradient in both relative and absolute risk, despite all included results being classified as negative. By providing absolute risks within the Australian screening setting, the study offers evidence directly relevant to evaluating whether participants with different negative f-Hb concentrations could benefit from different screening intervals. Implications of all the available evidence
A negative FIT result provides reassurance, but does not indicate the same level of interval CRC risk for every participant. Together, the international evidence and our findings support evaluating screening strategies that use quantitative f-Hb rather than relying solely on a positive or negative result in the Australian program. They do not establish which screening intervals or thresholds should be adopted. Further evaluation should determine whether tailoring screening reduces interval cancers and advanced disease without disproportionate increases in colonoscopy use, harms, or inequalities in access and outcomes.

## Introduction

Colorectal cancer (CRC) is the fifth most commonly diagnosed cancer and second leading cause of cancer death in Australia.^1^ It is highly preventable through polyp removal and highly treatable if detected early, with five-year relative survival of 99% for stage I disease.^2^ Australians have access to bowel cancer screening through the National Bowel Cancer Screening Program (NBCSP). Introduced in 2006 and fully implemented by 2020, the current NBCSP offers free biennial screening (every two years) to people aged 50 to 74 using a two-sample quantitative faecal immunochemical test (FIT) for the detection of occult blood. Since July 2024, people aged 45-49 years also became eligible for the NBCSP via an opt-in model, whereby they can request a free screening kit from the program.^3^

In cancer screening programs, interval cancers - i.e., cancers detected after a negative screening episode within the interval before the next episode - represent an important quality indicator of the program’s effectiveness and screening test sensitivity. In the NBCSP, this interval is limited to 24 months after a negative FIT. The reported evidence on interval CRC within the NBCSP is limited. The program’s latest monitoring report identified 701 interval CRCs among 1.1 million people with a negative or inconclusive screening result.^3^ However, this estimate was based on the 2018 screening data, before completion of the program’s biennial rollout to all eligible ages. Currently there are no contemporary estimates of the burden of interval CRC and how this burden may differ by participant characteristics, geography, tumour site, and the faecal heamoglobin (f-HB) concentration detected by the quantitative FIT values.

The Australian NBCSP uses a two-sample FIT protocol, with a positive result defined as a f-HB concentration of ≥20 µg of haemoglobin per gram of faeces (µg/g) in at least one of the two samples. FIT is often implemented in this way, as a qualitative test with a binary result (positive or negative) even though the test provides a quantitative f-Hb concentration. International studies have shown that f-Hb concentrations below a program’s positivity threshold are associated with subsequent CRC or colorectal neoplasia.^4,5^ Evidence on this relationship in the Australian context remains limited. Local evidence is particularly important because the two-sample protocol and 20 µg/g positivity threshold differ from those of international programs using a single sample or a higher threshold. Differences in f-Hb distributions and interval CRC risk by sex and age may also be relevant to precision-based screening, and therefore require evaluation within the local program.

In this study we used linked individual-level NBCSP and cancer registry data to provide a contemporary evaluation of interval CRC in the Australian context. Our study had two aims: 1) to estimate the burden, timing and cancer-site distribution of interval CRC; and 2) to quantify the association between f-Hb concentration and interval CRC by age and sex in the NBCSP.

## Methods

### Study design and data sources

We conducted a population-based retrospective cohort study using NBCSP screening records linked to the Queensland Cancer Registry, a population registry recording all cancers diagnosed in the state. Queensland is a state of Australia with a population of 5.7 million people (approximately 20% of the Australian population).^6^. The study population included all Queensland residents who returned a completed NBCSP FIT kit during a five-year period from 1 January 2018 to 31 December 2022. The dataset contained the individual-level f-HB results (in µg/g) for both samples for every screening round completed by each person, and all CRCs diagnosed during that time for each individual, as recorded in the Queensland Cancer Registry.

We used the screening episode as the unit of analysis, with the results from two stool samples. Participants could contribute more than one episode during 2018-2022, and analyses accounted for dependence between episodes from the same person (see below). Eligible episodes had numerical f-Hb concentrations below 20 µg/g in both samples - i.e., the NBCSP threshold for a positive result. We excluded episodes if either sample concentration was missing, the participant was outside the age range of 50-74 years, the earliest linked CRC diagnosis occurred on or before the start of follow-up, or dates were missing or invalid.

### Outcomes and follow-up

Interval CRC was the primary outcome and was defined as a CRC diagnosed within 24 months of a quantitative-negative screening episode and before the next completed NBCSP screening episode. Follow-up began on the latest of the two sample collection dates and the reported results date. It ended at the earliest of CRC diagnosis, the next completed NBCSP screening episode, death, 24 months after the FIT result date, or 31 December 2023. The next episode’s completion date was the later of its two sample collection dates. Person-time was calculated in days and converted to person-years. Tumour site was classified as right colon (ICD-O C18.0, C18.2-C18.4), left colon (C18.5-C18.7), rectosigmoid or rectum (C19.9, C20.9, with C21.8 retained in this group), or colon not otherwise specified (C18.9).

### Covariates

Pre-chosen covariates were age group, recorded sex, residential remoteness, socioeconomic disadvantage, and screening year. Age was grouped in five-year categories: 50-54, 55-59, 60-64, 65-69, and 70-74 years. Remoteness was classified using the Australian Statistical Geography Standard Remoteness Area, derived from ARIA+, and grouped as major cities, inner or outer regional, and remote or very remote.^7^ Area-level socioeconomic disadvantage was measured using quintiles of the Index of Relative Socio-economic Disadvantage (IRSD), assigned using residential postcode, with Q1 representing the most disadvantaged areas and Q5 the least disadvantaged.^8^ Screening year was included to account for differences between calendar-year screening cohorts.

For the quantitative f-Hb analyses, the main exposure variable was the maximum f-Hb concentration of the two samples in each screening episode, which reflects the NBCSP positivity criteria. Exploratory inspection of the data identified a marked change in the distribution of recorded sub-threshold concentrations before June 2021 (Figure S1). As the reason for this change could not be confirmed, we took a cautious approach and restricted concentration-specific analyses to episodes completed between 1 June 2021 and 31 December 2022, to reduce the potential influence of changes in laboratory measurement or NBCSP reporting.

### Statistical analysis

In the primary analysis, we used Cox proportional hazards models with time since FIT result as the time scale to estimate cause-specific hazard ratios (HRs) for interval CRC after a negative screening episode. To account for repeated screening by the same individual, we used Huber-White robust variance estimation^9^, with screening episodes clustered by participant. All statistical tests were two-sided. The adjusted model included age group, sex, remoteness, IRSD quintile, and FIT year.

We fitted Poisson models with a log person-time offset and the same covariates and participant clustering to estimate adjusted incidence rates, standardised to the complete-case cohort’s covariate and person-time distribution. Within the quantitative-analysis cohort, maximum f-Hb was analysed in four categories: 0-5 µg/g, >5-10 µg/g, >10-15 µg/g, and >15-<20 µg/g, with 0-5 µg/g as the reference. Continuous maximum f-Hb was represented by a natural cubic spline with four degrees of freedom, internal knots at 0.8, 3.8 and 11.8 µg/g, and boundary knots at 0 and 19.8 µg/g. The spline HR curve was expressed relative to 0 µg/g.

To provide absolute-risk estimates, we estimated 24-month cumulative incidence using the Aalen-Johansen estimator, treating death and completion of the next screening episode as competing events. These estimates describe the probability of an interval CRC before another screen or death under observed screening practice.

We described maximum f-Hb distributions by five-year age group and sex in the quantitative cohort. Age- and sex-specific 24-month cumulative incidence was estimated in both the full and complete-case quantitative cohorts. In the quantitative cohort, associations of age and sex with interval CRC were also estimated with adjustment for continuous f-Hb and the other covariates. Cumulative incidence by f-Hb category was estimated overall and separately for males and females.

## Ethics

Ethics approval was granted by the University of Melbourne Human Research Ethics Committee (Project ID: 28375).

## Results

### Cohort characteristics

The primary cohort included 1,083,149 quantitative-negative screening episodes from 700,734 participants, who contributed an average of 1.55 episodes each and up to five episodes over the study period (Figure S2). Just over half of the episodes were contributed by females, around four-fifths were from participants aged 50-69 years, and most were from people living in major cities (Table 1). The quantitative-analysis cohort included 326,977 episodes from June 2021 onwards, of which 326,931 had complete covariate data.

**Table 1.** Characteristics of the quantitative-negative screening cohort.

| **Characteristic** | **Negative episodes**<br>**n (%)** | **Interval CRCs**<br>**n (%)** |
| --- | --- | --- |
| All screening episodes | 1,083,149 | 586 |
| **Age at screening** |  |  |
| 50-54 | 230,855 (21.3) | 61 (10.4) |
| 55-59 | 173,294 (16.0) | 56 (9.6) |
| 60-64 | 266,701 (24.6) | 129 (22.0) |
| 65-69 | 182,855 (16.9) | 121 (20.6) |
| 70-74 | 229,444 (21.2) | 219 (37.4) |
| **Sex** |  |  |
| Female | 573,879 (53.0) | 273 (46.6) |
| Male | 509,270 (47.0) | 313 (53.4) |
| **Residential remoteness** |  |  |
| Major cities | 668,935 (61.8) | 316 (53.9) |
| Inner or outer regional | 400,960 (37.0) | 261 (44.5) |
| Remote or very remote | 13,231 (1.2) | 9 (1.5) |
| **IRSD quintile** |  |  |
| Q1 (most disadvantaged) | 238,313 (22.0) | 174 (29.7) |
| Q2 | 197,771 (18.3) | 103 (17.6) |
| Q3 | 262,225 (24.2) | 136 (23.2) |
| Q4 | 247,487 (22.8) | 111 (18.9) |
| Q5 (least disadvantaged) | 137,208 (12.7) | 62 (10.6) |
| **Screening year** |  |  |
| 2018 | 213,898 (19.7) | 152 (25.9) |
| 2019 | 247,785 (22.9) | 137 (23.4) |
| 2020 | 207,819 (19.2) | 105 (17.9) |
| 2021 | 225,176 (20.8) | 127 (21.7) |
| 2022 | 188,471 (17.4) | 65 (11.1) |
| **Maximum f-Hb, µg/g (quantitative cohort only)** |  |  |
| 0-5 | 293,921 (89.9) | 82 (59.4) |
| >5-10 | 20,789 (6.4) | 23 (16.7) |
| >10-15 | 7,826 (2.4) | 13 (9.4) |
| >15-<20 | 4,441 (1.4) | 20 (14.5) |

Remoteness was missing for 23 episodes and IRSD for 145, including the 23 with missing remoteness; none had an interval CRC. Percentages may not sum to 100 because of rounding or missing values. f-Hb is the maximum of the two samples. IRSD, Index of Relative Socio-economic Disadvantage.

### Interval CRC incidence and participant characteristics

Across 2,056,405 person-years of follow-up, 586 interval CRCs were diagnosed. The crude incidence was 28.5 per 100,000 person-years, and the estimated 24-month cumulative incidence was 57.6 per 100,000 negative screening episodes (95% CI 53.1-62.5; Table 2). After adjustment, people living in inner or outer regional areas had a higher interval CRC hazard than those living in major cities (HR 1.22, 95% CI 1.01-1.47). There was no consistent socioeconomic gradient. Although the hazard was lower in IRSD quintiles 2 and 4 than in the most disadvantaged quintile, it did not decrease progressively with decreasing disadvantage (Table 2).

**Table 2.** Interval CRC burden and adjusted associations in the full negative-screening cohort.

| Characteristic | CRC cases | Person-years | 24-month cumulative risk per 100,000 screening episodes (95% CI) | Adjusted incidence rate per 100,000 person-years (95% CI) | Adjusted HR (95% CI) | P value |
| --- | --- | --- | --- | --- | --- | --- |
| **Age at screening** |  |  |  |  |  |  |
| 50-54 | 61 | 437,788 | 28.0 (21.8-36.1) | 14.4 (11.2-18.5) | 1.00 (reference) | - |
| 55-59 | 56 | 325,562 | 35.0 (26.9-45.5) | 17.6 (13.5-22.9) | 1.22 (0.85-1.76) | 0.275 |
| 60-64 | 129 | 508,804 | 51.2 (43.1-60.9) | 25.1 (21.1-29.9) | 1.75 (1.29-2.37) | <0.001 |
| 65-69 | 121 | 348,218 | 69.9 (58.5-83.6) | 34.1 (28.5-40.8) | 2.37 (1.74-3.23) | <0.001 |
| 70-74 | 219 | 436,033 | 101.4 (88.8-115.8) | 49.1 (43.0-56.0) | 3.42 (2.57-4.54) | <0.001 |
| **Sex** |  |  |  |  |  |  |
| Female | 273 | 1,091,472 | 50.7 (45.0-57.1) | 25.2 (22.4-28.4) | 1.00 (reference) | - |
| Male | 313 | 964,933 | 65.4 (58.5-73.1) | 32.2 (28.8-35.9) | 1.28 (1.08-1.50) | 0.003 |
| **Residential remoteness** |  |  |  |  |  |  |
| Major cities | 316 | 1,269,327 | 50.2 (45.0-56.1) | 26.1 (23.2-29.3) | 1.00 (reference) | - |
| Inner or outer regional | 261 | 761,694 | 69.5 (61.5-78.5) | 31.8 (27.9-36.3) | 1.22 (1.01-1.47) | 0.038 |
| Remote or very remote | 9 | 25,339 | 70.8 (36.8-136.2) | 35.5 (18.4-68.6) | 1.36 (0.70-2.66) | 0.368 |
| **IRSD quintile** |  |  |  |  |  |  |
| Q1 (most disadvantaged) | 174 | 452,737 | 77.9 (67.1-90.4) | 34.7 (29.7-40.6) | 1.00 (reference) | - |
| Q2 | 103 | 376,068 | 55.4 (45.6-67.2) | 25.6 (21.0-31.2) | 0.74 (0.58-0.94) | 0.014 |
| Q3 | 136 | 497,988 | 54.9 (46.4-65.0) | 27.7 (23.4-32.8) | 0.80 (0.63-1.00) | 0.055 |
| Q4 | 111 | 469,577 | 48.0 (39.8-57.8) | 25.5 (21.0-31.0) | 0.73 (0.57-0.95) | 0.019 |
| Q5 (least disadvantaged) | 62 | 259,764 | 48.1 (37.5-61.7) | 27.3 (21.1-35.3) | 0.79 (0.58-1.07) | 0.129 |

Adjusted Cox HRs and Poisson rates use 1,083,004 complete-case episodes and adjust for all listed covariates and screening year. Poisson rates are standardised to the cohort's covariate and person-time distribution. Confidence intervals account for repeated episodes within participants. HR, hazard ratio; CI, confidence interval; IRSD, Index of Relative Socio-economic Disadvantage. P values are two-sided participant-robust Wald tests for individual Cox model contrasts with the reference group.

### Timing and tumour site

Interval CRC diagnoses were more frequent in the second year after screening than in the first. The number of diagnoses increased across successive six-month periods, from 103 within the first six months to 188 between 18 and 24 months (Table S6; Figure S3). Almost half of the cancers arose in the right colon (283; 48.3%), followed by the rectosigmoid or rectum (200; 34.1%) and the left colon (94; 16.0%). The remaining nine cancers had no specific colonic site recorded.

### Faecal haemoglobin concentration and interval CRC risk

In the complete-case quantitative cohort, 138 interval CRCs occurred during 554,720 person-years of follow-up. Risk increased substantially across the negative f-Hb range (Table 3). Compared with a maximum f-Hb concentration of 0-5 µg/g, the adjusted hazard was higher in each successive category, reaching an HR of 14.93 (95% CI 9.02-24.70) at >15-<20 µg/g. The continuous analysis showed the same increasing risk across the negative range (Figure 1).

**Figure 1.**
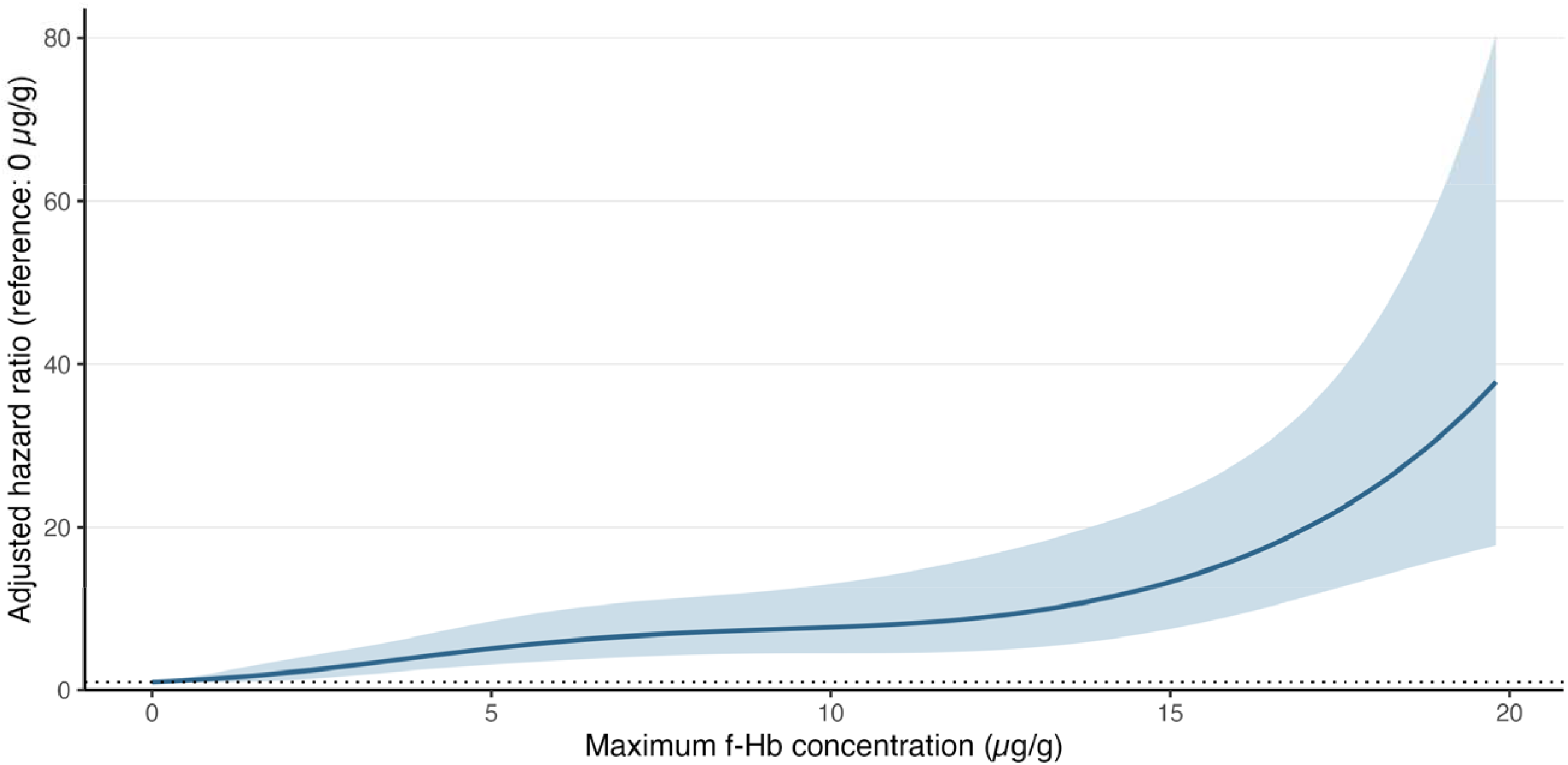
Interval CRC hazard across the negative f-Hb range. The spline model has four degrees of freedom and adjusts for five-year age group, sex, remoteness, IRSD quintile and screening year, with participant-clustered robust variances.

**Table 3.** Quantitative f-Hb concentration and interval CRC risk.

| Maximum f-Hb (µg/g) | Screening episodes | CRC cases | Person-years | 24-month cumulative risk per 100,000 screening episodes (95% CI) | Adjusted incidence rate per 100,000 person-years (95% CI) | Adjusted HR (95% CI) | P value |
| --- | --- | --- | --- | --- | --- | --- | --- |
| 0-5 | 293,876 | 82 | 497,297 | 35.7 (28.5-44.7) | 16.6 (13.4-20.6) | 1.00 (reference) | - |
| >5-10 | 20,788 | 23 | 36,273 | 132.1 (87.1-200.3) | 60.5 (40.1-91.3) | 3.65 (2.29-5.82) | <0.001 |
| >10-15 | 7,826 | 13 | 13,517 | 182.1 (105.4-314.6) | 90.9 (52.6-156.9) | 5.48 (3.04-9.90) | <0.001 |
| >15-<20 | 4,441 | 20 | 7,633 | 504.5 (323.4-787.0) | 247.7 (157.9-388.6) | 14.93 (9.02-24.70) | <0.001 |

The quantitative-analysis cohort includes episodes from 1 June 2021 to 31 December 2022. All columns use the same 326,931 complete-case episodes (138 cancers). Maximum f-Hb is the higher of the two sample values. CI, confidence interval; HR, hazard ratio. P values are two-sided participant-robust Wald tests for individual Cox model contrasts with the reference group.

The gradient was also evident in absolute risk. Observed 24-month cumulative incidence increased from 35.7 per 100,000 screening episodes (95% CI 28.5-44.7) in the lowest category to 504.5 (323.4-787.0) in the highest. Adjusted incidence rates showed a similar pattern (Table 3).

### Age differences

Interval CRC risk increased with age in the full cohort and was highest among participants aged 70-74 years. This association remained after adjustment for sex, remoteness, socioeconomic disadvantage and screening year (Table 2; Figure 2). In the quantitative cohort, participants aged 65-69 and 70-74 years also had higher risk than those aged 50-54 years after adjustment for f-Hb and the other covariates. Higher negative-range f-Hb concentrations became more common with increasing age in both males and females (Figure S5).

**Figure 2.**
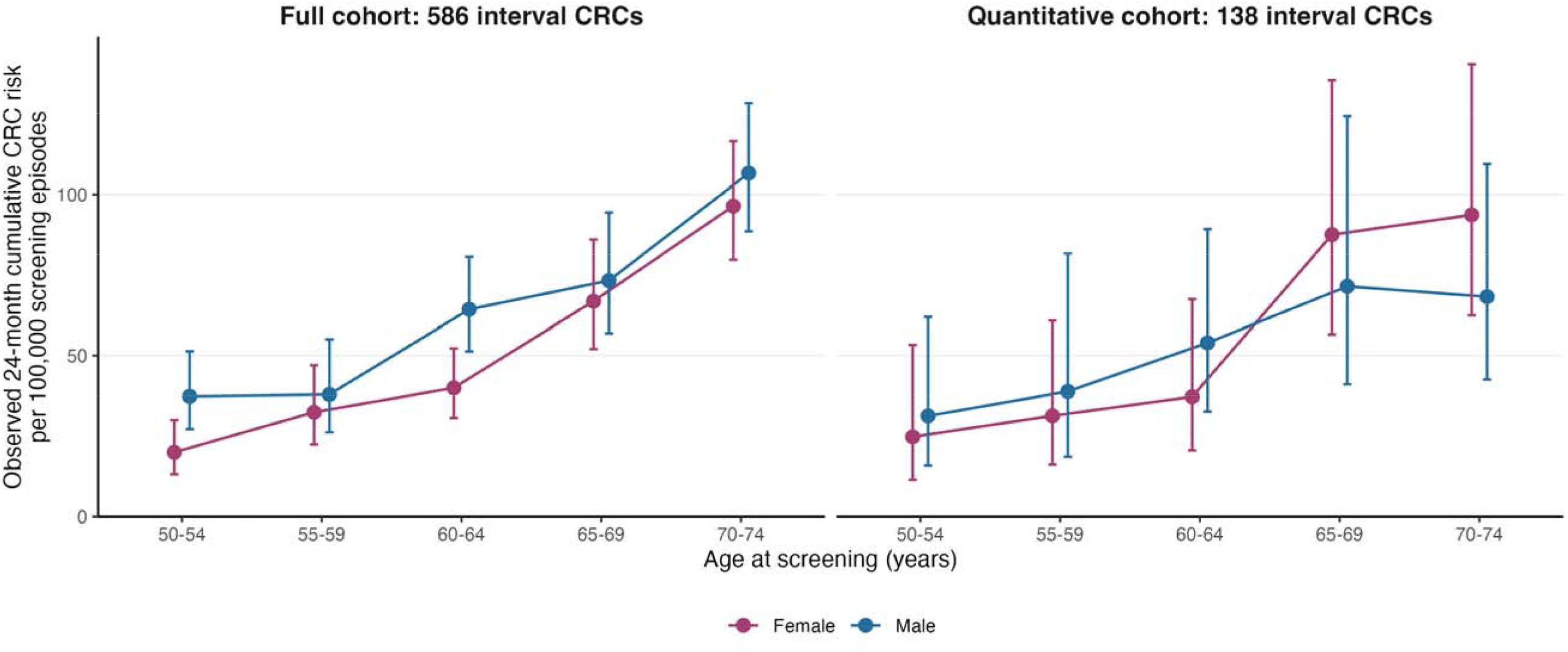
Observed 24-month interval CRC risk by age group and sex. Observed Aalen-Johansen cumulative incidence (%) with participant-clustered 95% CIs; death and the next screen are competing events. Full cohort: 1,083,149 episodes and 586 CRCs. Quantitative cohort: 326,931 episodes and 138 CRCs. Estimates are unadjusted; counts and risks are presented in Table S5.

### Sex differences

In the full cohort, males had higher interval CRC risk than females, including after adjustment for age and the other covariates (Table 2; Figure 2). In the smaller quantitative cohort, however, observed risks were similar for males and females (Table S5). The analysis adjusted for continuous f-Hb and the other covariates also provided no clear evidence of a sex difference, although the estimate was imprecise.

Males were more likely than females to have f-Hb concentrations above 5 µg/g in every age group. They also had a higher proportion of results recorded as zero, so the difference between the sexes was not a uniform shift towards higher concentrations. Some combinations of age, sex and f-Hb category contained few or no interval CRCs (Table S1).

### Cumulative incidence and reference-risk comparisons

Cumulative incidence rose more rapidly at higher f-Hb concentrations (Figure 3). The 24-month risk in the 0-5 µg/g group was reached at approximately 10.4 months in the >5-10 µg/g group, 5.4 months in the >10-15 µg/g group and 0.4 months in the >15-<20 µg/g group. Corresponding model-standardised times were 10.1, 6.6 and 2.7 months (Figure S4; Table S4).

**Figure 3.**
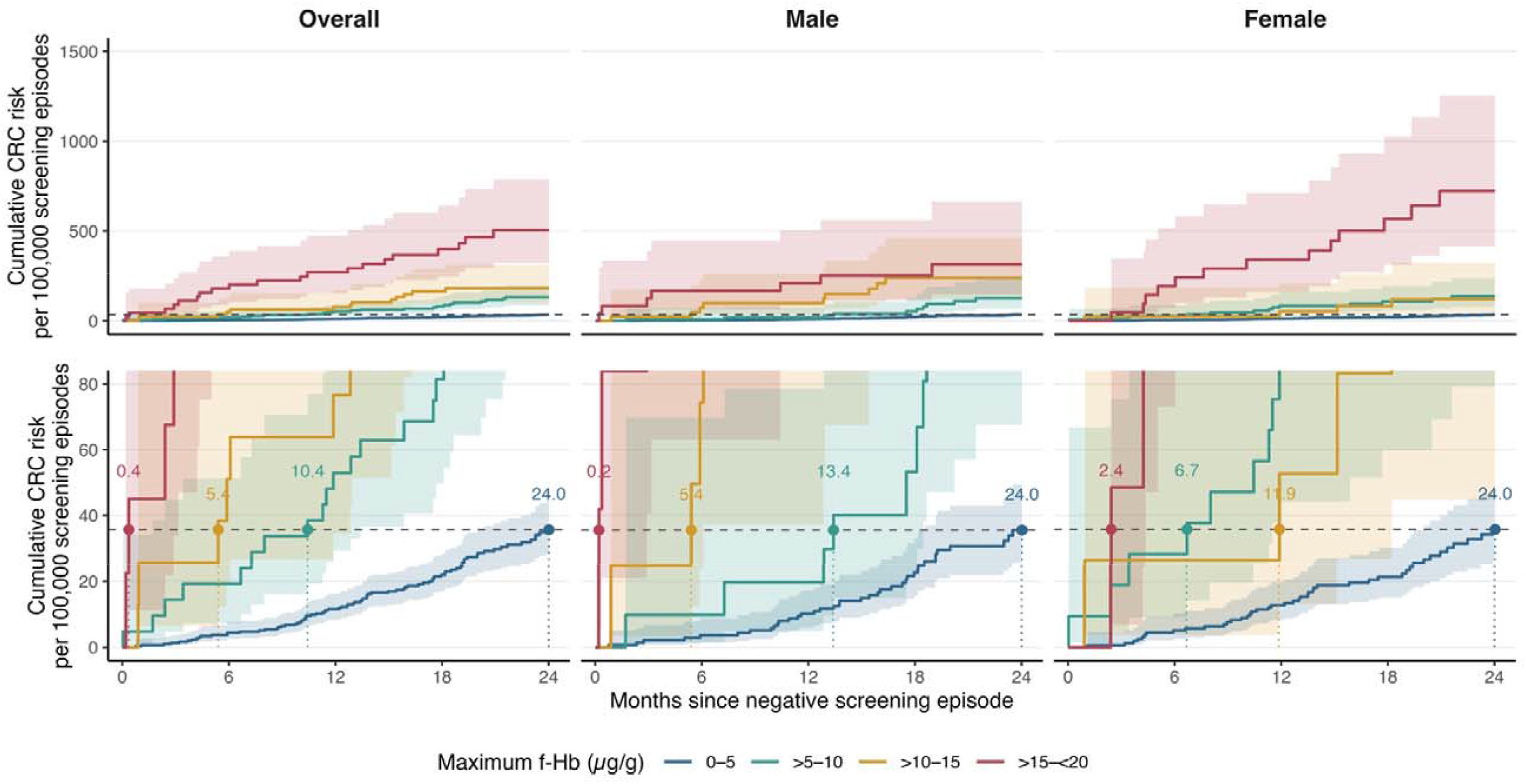
Observed cumulative incidence by f-Hb category and time to a common reference risk. Aalen-Johansen cumulative incidence overall and by sex in the quantitative-analysis cohort. Upper panels show the full concentration gradient; lower panels enlarge the low-risk region. The dashed horizontal line mark the 24-month risk for 0-5 µg/g. Coloured points and labels show the first crossing time in months Curves and crossings are point estimates and illustrate risk accumulation under observed screening practice. Endpoint confidence intervals and numbers at risk are presented in Tables S2-S3

The very early crossing in the highest category occurred at the second diagnosis in that group. Sex-specific curves showed the same broad concentration gradient, but small event counts produced irregular early steps. These comparisons describe when groups reached a common level of risk; they do not establish when repeat FIT would prevent or detect a cancer.

## Discussion

In this large linked Queensland NBCSP cohort, interval CRC after a negative screening episode was uncommon but varied substantially with the recorded f-Hb concentration. Across more than 1.08 million negative screening episodes, the cumulative interval CRC incidence was 57.6 per 100,000 screening episodes. This equates to approximately 130 diagnoses of CRC over the following two years among the ∼ 224,000 Queensland residents who participate each year in the NBCSP but test negative. In the quantitative cohort, cumulative incidence increased approximately 14-fold, from 36 per 100,000 screening episodes at 0-5 µg/g to 504 per 100,000 at >15-<20 µg/g, with an adjusted HR of 14.93 for the highest versus lowest category. These findings show that quantitative f-Hb provides useful prognostic information beyond the binary classification of FIT results as positive or negative. Although absolute risk was low for most participants, the variation within the negative range provides a basis for evaluating more personalised screening.

Our findings are broadly consistent with the limited Australian evidence. Our overall interval CRC risk estimate was close to the 63 per 100,000 people reported by the NBCSP based on the 2018 screening cohort.^3^ A study in Western Australia, based on the same 2018 data and including 122,851 NBCSP participants identified 51 interval CRCs and reported higher risk among males and older participants.^10^ Our study extends these findings using screening records from 2018-2022, covering 700,734 NBCSP participants in Queensland and 586 interval CRCs providing more recent evidence from a larger state cohort. Our analysis is the first to show how interval CRC risk varies across the negative f-Hb range in the NBCSP’s two-sample protocol.

Internationally, multiple studies have described interval CRC after negative stool testing. Wieten and colleagues reviewed 29 studies, 17 of which used FIT, and estimated a pooled interval CRC incidence of 20 (95% CI 14-29) per 100,000 person-years after negative FIT.^11^ The authors identified substantial heterogeneity between studies, which means that their pooled estimate should not be interpreted as a benchmark for FIT screening performance. Within this literature, a smaller group of studies examined interval CRC across the negative f-Hb range. In Denmark, where the positivity threshold is the same as in Australia (20 µg/g), Deding and colleagues. found increasing interval CRC risk across the negative f-Hb range (HR of 7.09 for f-Hb concentrations of 11.9-19.8 µg/g compared with <7.2 µg/g).^12^ In the Dutch program, which uses a 47 µg/g threshold, Breekveldt and colleagues found a concentration gradient after both the first and second negative tests (Odds Ratio of 7.2 for concentrations of >10-20 µg/g compared with 0-2.6 µg/g after both the first and second negative tests.^13^ These findings support interpreting negative FIT results as a range of risks rather than a uniform low-risk group. This prognostic interpretation is consistent with international evidence beyond interval CRC. A recent dose-response meta-analysis of 13 studies linked higher negative-range f-Hb to subsequent colorectal neoplasia.^5^

Interval CRC is important despite its low absolute incidence. In Taiwan, Hsu and colleagues identified 2,782 post-FIT interval CRCs in the national screening program, accounting for 18.1% of all CRCs diagnosed in the program.^14^ Tran and colleagues found that 1,534 interval cancers in Flanders were more often right-sided, advanced-stage and high-grade than screen-detected cancers.^15^ In our cohort, 283 of the 586 interval CRCs were right-sided (48.3%). This is consistent with the data from Western Australia where 25 of 51 interval CRCs were right-sided. A Swedish studies found a similar anatomical distribution in the population-based regional program of Stockholm-Gotland where 48.4% of FIT-negative interval cancers were proximal.^16^

Our analysis showed that older participants had higher interval CRC risk and more upper-range negative f-Hb results. Analysis of Danish program also found an age gradient after adjustment for f-Hb.^12^ Among Dutch participants reaching the screening upper age limit, van Stigt and colleagues found that detectable f-Hb at the final negative test was associated with approximately fivefold interval CRC risk.^17^ These studies along with our analysis highlight the importance of age when interpreting risk after a negative result.

Males had a higher risk of interval CRC in our full cohort, whereas no clear male excess was evident in the smaller quantitative cohort. The latter estimate was imprecise, with overlapping confidence intervals compatible with both lower and higher risk in males. Therefore, it does not establish equal risks or explain the full-cohort association. In Denmark, a male excess interval CRC risk persisted after adjustment for f-Hb.^12^ In Scotland, Clark and colleagues reported a higher interval cancer proportion among females, indicating lower FIT sensitivity higher interval-cancer proportion among women has been reported.^18^ However, that proportion uses all diagnosed cancers as its denominator, which differs from the absolute risk of interval CRC after a negative FIT examined in our study.

The concentration gradient provides a rationale for evaluating screening intervals that take f-Hb concentration into account.

In a recent study, Yen and colleagues used data from 3.5 million participants in Taiwan’s screening program to model screening intervals tailored to f-Hb concentration, with shorter intervals for higher concentrations and longer intervals for lower concentrations.(19) The intervals were designed to preserve the effectiveness of screening everyone every two years. Under this modelling assumption, the strategy was projected to require 49% fewer FITs and 28% fewer colonoscopies, assuming an 80% colonoscopy referral rate after a positive FIT.^19^ Our cumulative-incidence curves provide a complementary view within the current two-year interval window in Australia: groups with higher f-Hb concentration groups reached the lowest category’s two-year risk earlier. This pattern supports evaluating earlier repeat testing for people with higher negative-range concentrations. The Taiwanese modelling also highlights the potential to redistribute screening resources according to risk, an approach that warrants assessment within the NBCSP.

Importantly, any such strategy must be evaluated in terms of cancer outcomes, harms, participation and resource use. Dutch screening data showed higher CRC sensitivity at a lower FIT threshold,^20^ while a systematic review of studies using colonoscopy to verify FIT results found that lower thresholds detected more cancers but also produced more positive results among people without CRC.^21^ Lower thresholds or more frequent testing may increase the demand for colonoscopy and the number of false-positive results. Simulation modelling studies can be used to assess the potential benefits, harms and cost-effectiveness of strategies informed by these quantitative risk estimates. Any added complexity to screening eligibility and process must preserve participation in initial testing, repeat screening and colonoscopy after a positive result.^22^ The higher adjusted risk in regional residents and differences between some IRSD quintiles seen in our analyses also make equitable access to investigation and follow-up an important consideration for implementation.

The main strengths of this study include its population-based linkage, a large cohort, and accounting for repeated screening records. We accounted for repeated episodes within participants and for death and subsequent screening as competing events, providing estimates of risk under observed program practice. Several differences nevertheless limit direct numerical comparisons with other studies. The analysis presented in the NBCSP monitoring report includes negative or inconclusive results, and the Western Australian study could not end follow-up at the next screen, which means that cancers detected at another screening round within 24 months could have been classified as interval cancers.^10^ Our definition used f-HB values below 20 µg/g and only included cancers diagnosed before the next completed screen.

Measurement quality is an important limitation. Concentration analyses were restricted to June 2021 onwards because earlier records showed a marked change in low f-Hb values. Laboratory confirmation of the cause and validity of the retained measurements was unavailable. Detection limits, handling of low values and reporting changes can affect recorded concentrations, while assay performance can differ at the same nominal threshold.^23^ These issues require verification before concentrations are used to guide screening frequency and recall within the NBCSP.

The quantitative cohort contained only 138 interval CRCs, limiting precision in some age, sex and concentration groups. Later episodes had shorter follow-up, and estimates assume non-informative administrative censoring. Findings from Queensland participants may not generalise to other jurisdictions. We had no polyp data and could not assess broader colorectal neoplasia outcomes. Cancer-stage data were also unavailable, so we could not determine whether higher-risk groups were more likely to have advanced-stage disease.

In conclusion, quantitative f-Hb below the NBCSP positivity threshold identifies substantial differences in interval CRC risk. Retaining this information and assessing it alongside age and sex could support more personalised screening. Laboratory verification, further validation of our estimates, and prospective assessment of benefits, harms and feasibility are needed before changing recall intervals or thresholds.

## Data Availability

Data analyses in the present work are available upon request/application to the data custodians: Queensland Health and the Queensland Cancer Registry.

