## Supplementary material for "Interval colorectal cancer risk by haemoglobin concentration in Australia’s National Bowel Cancer Screening Program"

Supplementary appendix

**Table S1. Interval CRC counts and crude rates by age group, sex, and f-Hb**

| **Age** | **Sex** | **Maximum f-Hb (µg/g)** | **Negative episodes** | **Interval CRCs** | **Person-years** | **Crude rate** |
| --- | --- | --- | --- | --- | --- | --- |
| 50–54 | Female | 0–5 | 36,512 | 2 | 61,493 | 3.3 |
|  |  | >5–10 | 2,127 | 2 | 3,728 | 53.6 |
|  |  | >10–15 | 725 | 0 | 1,247 | 0.0 |
|  |  | >15–<20 | 407 | 3 | 700 | 428.3 |
| 50–54 | Male | 0–5 | 31,408 | 4 | 53,585 | 7.5 |
|  |  | >5–10 | 1,908 | 1 | 3,313 | 30.2 |
|  |  | >10–15 | 731 | 2 | 1,271 | 157.4 |
|  |  | >15–<20 | 417 | 2 | 722 | 277.0 |
| 55–59 | Female | 0–5 | 29,093 | 6 | 48,473 | 12.4 |
|  |  | >5–10 | 1,715 | 2 | 2,955 | 67.7 |
|  |  | >10–15 | 616 | 0 | 1,045 | 0.0 |
|  |  | >15–<20 | 321 | 1 | 539 | 185.5 |
| 55–59 | Male | 0–5 | 25,297 | 5 | 42,133 | 11.9 |
|  |  | >5–10 | 1,566 | 1 | 2,667 | 37.5 |
|  |  | >10–15 | 603 | 2 | 1,025 | 195.1 |
|  |  | >15–<20 | 380 | 0 | 647 | 0.0 |
| 60–64 | Female | 0–5 | 37,180 | 6 | 63,337 | 9.5 |
|  |  | >5–10 | 2,538 | 3 | 4,498 | 66.7 |
|  |  | >10–15 | 883 | 0 | 1,549 | 0.0 |
|  |  | >15–<20 | 479 | 3 | 829 | 361.9 |
| 60–64 | Male | 0–5 | 31,741 | 10 | 54,057 | 18.5 |
|  |  | >5–10 | 2,395 | 1 | 4,195 | 23.8 |
|  |  | >10–15 | 985 | 2 | 1,709 | 117.1 |
|  |  | >15–<20 | 558 | 3 | 958 | 313.0 |
| 65–69 | Female | 0–5 | 24,773 | 10 | 42,123 | 23.7 |
|  |  | >5–10 | 1,881 | 3 | 3,326 | 90.2 |
|  |  | >10–15 | 666 | 3 | 1,156 | 259.6 |
|  |  | >15–<20 | 348 | 5 | 599 | 834.2 |
| 65–69 | Male | 0–5 | 21,529 | 8 | 36,622 | 21.8 |
|  |  | >5–10 | 1,807 | 4 | 3,144 | 127.2 |
|  |  | >10–15 | 760 | 1 | 1,317 | 75.9 |
|  |  | >15–<20 | 421 | 0 | 727 | 0.0 |
| 70–74 | Female | 0–5 | 29,320 | 20 | 49,756 | 40.2 |
|  |  | >5–10 | 2,353 | 3 | 4,143 | 72.4 |
|  |  | >10–15 | 903 | 1 | 1,569 | 63.7 |
|  |  | >15–<20 | 502 | 1 | 871 | 114.8 |
| 70–74 | Male | 0–5 | 27,023 | 11 | 45,718 | 24.1 |
|  |  | >5–10 | 2,498 | 3 | 4,304 | 69.7 |
|  |  | >10–15 | 954 | 2 | 1,632 | 122.5 |
|  |  | >15–<20 | 608 | 2 | 1,039 | 192.4 |

**Table S2. Observed cumulative incidence at 6, 12, 18, and 24 months**

| **Maximum f-Hb (µg/g)** | **6 months** | **12 months** | **18 months** | **24 months** |
| --- | --- | --- | --- | --- |
| **Overall** |  |  |  |  |
| 0–5 | 4.4 (2.6–7.6) | 11.6 (8.3–16.2) | 22.0 (17.0–28.3) | 35.7 (28.5–44.7) |
| >5–10 | 19.2 (7.2–51.3) | 52.9 (29.3–95.5) | 81.4 (49.7–133.2) | 132.1 (87.1–200.3) |
| >10–15 | 51.1 (19.2–136.1) | 76.7 (34.5–170.6) | 164.0 (93.0–289.3) | 182.1 (105.4–314.6) |
| >15–<20 | 180.1 (90.1–360.0) | 270.2 (153.6–475.4) | 399.1 (247.8–642.6) | 504.5 (323.4–787.0) |
| **Male** |  |  |  |  |
| 0–5 | 3.6 (1.5–8.8) | 10.2 (6.1–17.3) | 22.6 (15.6–32.9) | 35.6 (25.6–49.5) |
| >5–10 | 9.8 (1.4–69.8) | 19.7 (4.9–78.6) | 53.2 (22.0–128.7) | 127.0 (67.6–238.5) |
| >10–15 | 74.4 (24.0–230.5) | 99.2 (37.2–264.1) | 239.7 (124.5–461.5) | 239.7 (124.5–461.5) |
| >15–<20 | 167.8 (63.0–446.7) | 209.7 (87.4–503.4) | 252.7 (113.6–561.9) | 314.9 (148.9–665.8) |
| **Female** |  |  |  |  |
| 0–5 | 5.1 (2.6–10.2) | 12.7 (8.2–19.8) | 21.3 (15.1–30.2) | 35.8 (26.2–48.8) |
| >5–10 | 28.3 (9.1–87.6) | 84.8 (44.1–162.9) | 108.3 (59.8–196.0) | 137.6 (79.2–239.1) |
| >10–15 | 26.4 (3.7–187.1) | 52.7 (13.2–210.8) | 83.3 (26.8–258.8) | 120.5 (44.8–324.3) |
| >15–<20 | 194.5 (73.1–517.6) | 340.3 (162.4–712.9) | 568.0 (314.1–1027.2) | 722.8 (416.4–1254.8) |

**Table S3. Numbers at risk for the observed cumulative-incidence curves**

| **Maximum f-Hb (µg/g)** | **0 months** | **6 months** | **12 months** | **18 months** | **24 months** |
| --- | --- | --- | --- | --- | --- |
| **Overall** |  |  |  |  |  |
| 0–5 | 293,876 | 293,625 | 292,984 | 198,853 | 120,946 |
| >5–10 | 20,788 | 20,751 | 20,685 | 15,161 | 10,779 |
| >10–15 | 7,826 | 7,811 | 7,787 | 5,547 | 3,822 |
| >15–<20 | 4,441 | 4,420 | 4,398 | 3,126 | 2,092 |
| **Male** |  |  |  |  |  |
| 0–5 | 136,998 | 136,838 | 136,483 | 93,159 | 56,928 |
| >5–10 | 10,174 | 10,149 | 10,113 | 7,302 | 5,041 |
| >10–15 | 4,033 | 4,025 | 4,006 | 2,847 | 1,946 |
| >15–<20 | 2,384 | 2,373 | 2,360 | 1,672 | 1,116 |
| **Female** |  |  |  |  |  |
| 0–5 | 156,878 | 156,787 | 156,501 | 105,694 | 64,018 |
| >5–10 | 10,614 | 10,602 | 10,572 | 7,859 | 5,738 |
| >10–15 | 3,793 | 3,786 | 3,781 | 2,700 | 1,876 |
| >15–<20 | 2,057 | 2,047 | 2,038 | 1,454 | 976 |

**Table S4. Time to the panel-specific 24-month reference risk**

| **Estimate and group** | **Maximum f-Hb (µg/g)** | **Reference cumulative risk per 100,000 screening episodes** | **Crossing time months** | **24-month cumulative risk per 100,000 screening episodes** |
| --- | --- | --- | --- | --- |
| Observed: Overall | 0–5 | 35.7 | 24.0 | 35.7 |
|  | >5–10 | 35.7 | 10.4 | 132.1 |
|  | >10–15 | 35.7 | 5.4 | 182.1 |
|  | >15–<20 | 35.7 | 0.4 | 504.5 |
| Observed: Male | 0–5 | 35.6 | 24.0 | 35.6 |
|  | >5–10 | 35.6 | 13.4 | 127.0 |
|  | >10–15 | 35.6 | 5.4 | 239.7 |
|  | >15–<20 | 35.6 | 0.2 | 314.9 |
| Observed: Female | 0–5 | 35.8 | 24.0 | 35.8 |
|  | >5–10 | 35.8 | 6.7 | 137.6 |
|  | >10–15 | 35.8 | 11.9 | 120.5 |
|  | >15–<20 | 35.8 | 2.4 | 722.8 |
| Adjusted: Overall | 0–5 | 35.2 | 24.0 | 35.2 |
|  | >5–10 | 35.2 | 10.1 | 128.2 |
|  | >10–15 | 35.2 | 6.6 | 192.4 |
|  | >15–<20 | 35.2 | 2.7 | 521.7 |
| Adjusted: Male | 0–5 | 33.6 | 24.0 | 33.6 |
|  | >5–10 | 33.6 | 10.1 | 122.2 |
|  | >10–15 | 33.6 | 6.6 | 183.5 |
|  | >15–<20 | 33.6 | 2.7 | 497.3 |
| Adjusted: Female | 0–5 | 36.6 | 24.0 | 36.6 |
|  | >5–10 | 36.6 | 10.1 | 133.4 |
|  | >10–15 | 36.6 | 6.6 | 200.3 |
|  | >15–<20 | 36.6 | 2.7 | 543.4 |

**Table S5. Observed 24-month interval CRC risk by age group and sex**

| **Age** | **Sex** | **Negative episodes** | **Interval CRCs** | **Person-years** | **24-month cumulative risk per 100,000 screening episodes**  **(95% CI)** |
| --- | --- | --- | --- | --- | --- |
| **Full cohort** | | | | | |
| All ages | Female | 573,879 | 273 | 1,091,472 | 50.7 (45.0–57.1) |
|  | Male | 509,270 | 313 | 964,933 | 65.4 (58.5–73.1) |
| 50–54 | Female | 123,289 | 23 | 233,546 | 19.9 (13.2–30.0) |
|  | Male | 107,566 | 38 | 204,242 | 37.3 (27.1–51.3) |
| 55–59 | Female | 92,770 | 28 | 174,579 | 32.4 (22.3–47.0) |
|  | Male | 80,524 | 28 | 150,983 | 37.9 (26.2–55.0) |
| 60–64 | Female | 143,710 | 54 | 274,672 | 40.0 (30.6–52.2) |
|  | Male | 122,991 | 75 | 234,132 | 64.3 (51.3–80.7) |
| 65–69 | Female | 96,483 | 61 | 184,119 | 66.9 (52.0–86.0) |
|  | Male | 86,372 | 60 | 164,099 | 73.3 (56.9–94.4) |
| 70–74 | Female | 117,627 | 107 | 224,556 | 96.4 (79.8–116.6) |
|  | Male | 111,817 | 112 | 211,477 | 106.6 (88.6–128.3) |
| **Quantitative cohort** | | | | | |
| All ages | Female | 173,342 | 74 | 293,936 | 52.5 (41.4–66.6) |
|  | Male | 153,589 | 64 | 260,784 | 51.7 (40.2–66.6) |
| 50–54 | Female | 39,771 | 7 | 67,168 | 24.7 (11.5–53.3) |
|  | Male | 34,464 | 9 | 58,891 | 31.3 (15.8–62.0) |
| 55–59 | Female | 31,745 | 9 | 53,012 | 31.3 (16.1–60.9) |
|  | Male | 27,846 | 8 | 46,472 | 38.9 (18.5–81.6) |
| 60–64 | Female | 41,080 | 12 | 70,212 | 37.2 (20.5–67.5) |
|  | Male | 35,679 | 16 | 60,919 | 54.0 (32.6–89.2) |
| 65–69 | Female | 27,668 | 21 | 47,204 | 87.5 (56.5–135.5) |
|  | Male | 24,517 | 13 | 41,810 | 71.5 (41.1–124.4) |
| 70–74 | Female | 33,078 | 25 | 56,340 | 93.7 (62.5–140.5) |
|  | Male | 31,083 | 18 | 52,693 | 68.2 (42.6–109.4) |

**Table S6. Timing and anatomical site of interval CRC diagnoses**

| **Characteristic** | **Interval CRCs** | **%** |
| --- | --- | --- |
| **Time since negative screening episode** |  |  |
| 0–6 months | 103 | 17.6 |
| >6–12 months | 120 | 20.5 |
| >12–18 months | 175 | 29.9 |
| >18–24 months | 188 | 32.1 |
| **Anatomical site** |  |  |
| Right colon | 283 | 48.3 |
| Left colon | 94 | 16.0 |
| Rectosigmoid or rectum | 200 | 34.1 |
| Colon not otherwise specified | 9 | 1.5 |

**Figure S1. Monthly distribution of recorded faecal haemoglobin concentrations**

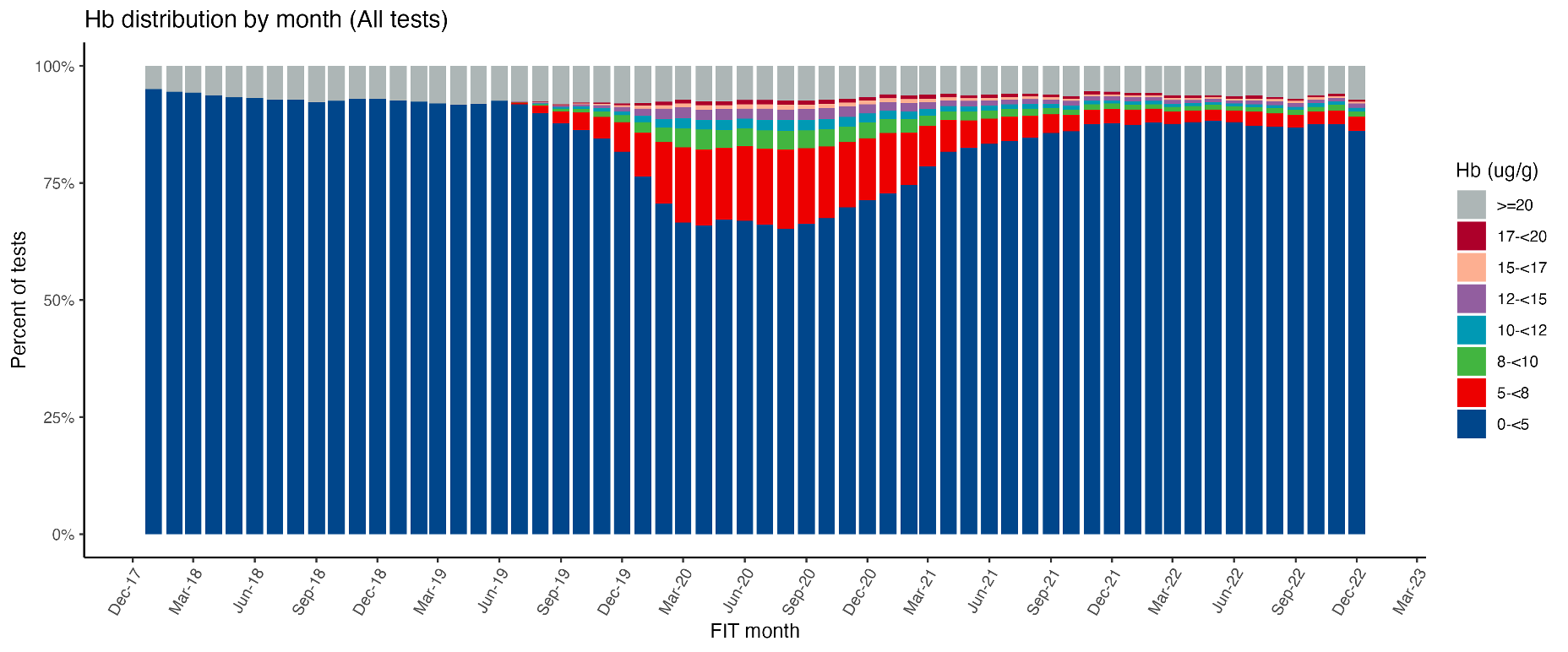

Stacked bars show the percentage of tests in each concentration range, including results at or above 20 µg/g.

**Figure S2. Derivation of the analysis cohorts**

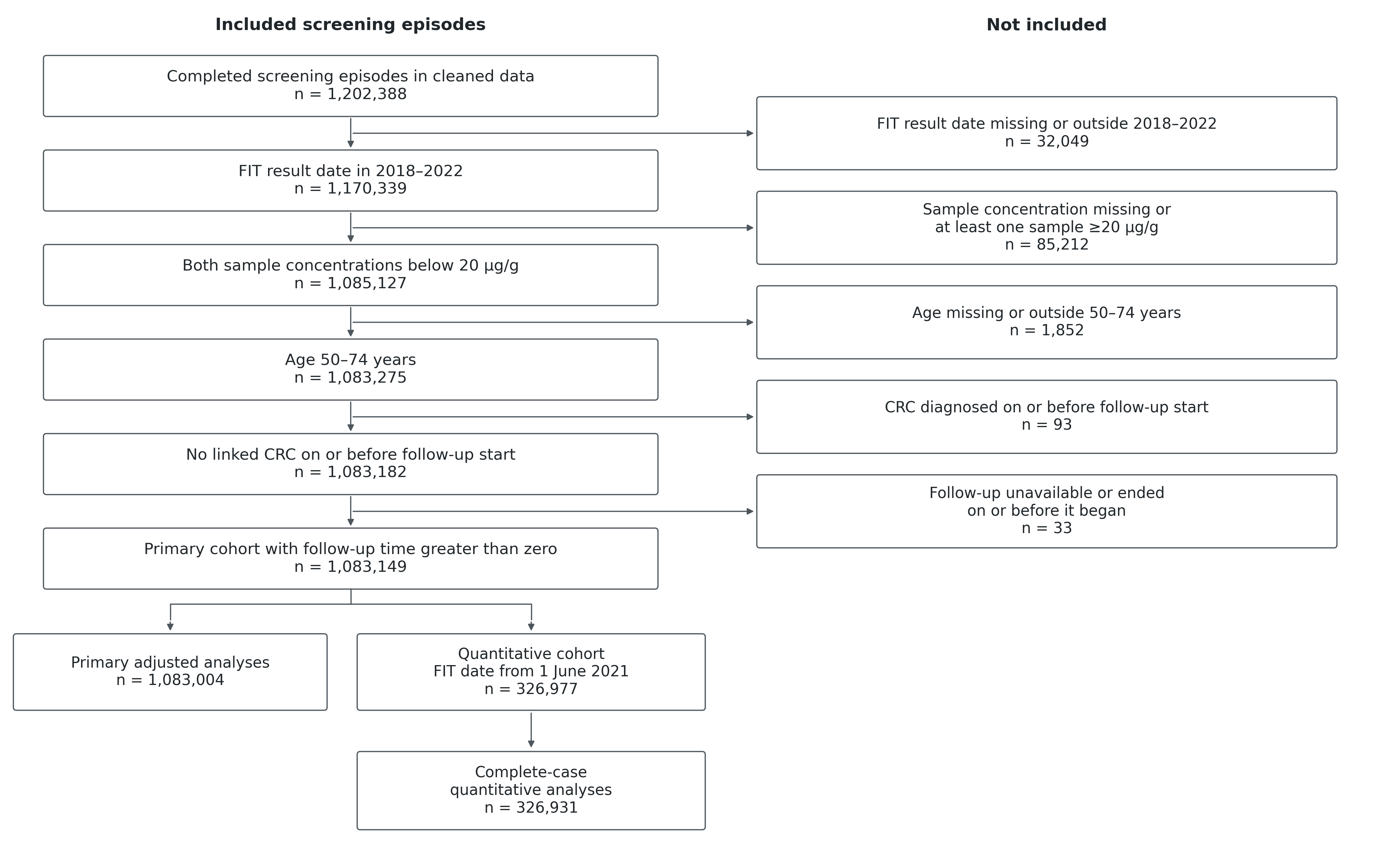

**Figure S3. Timing of interval CRC diagnoses**

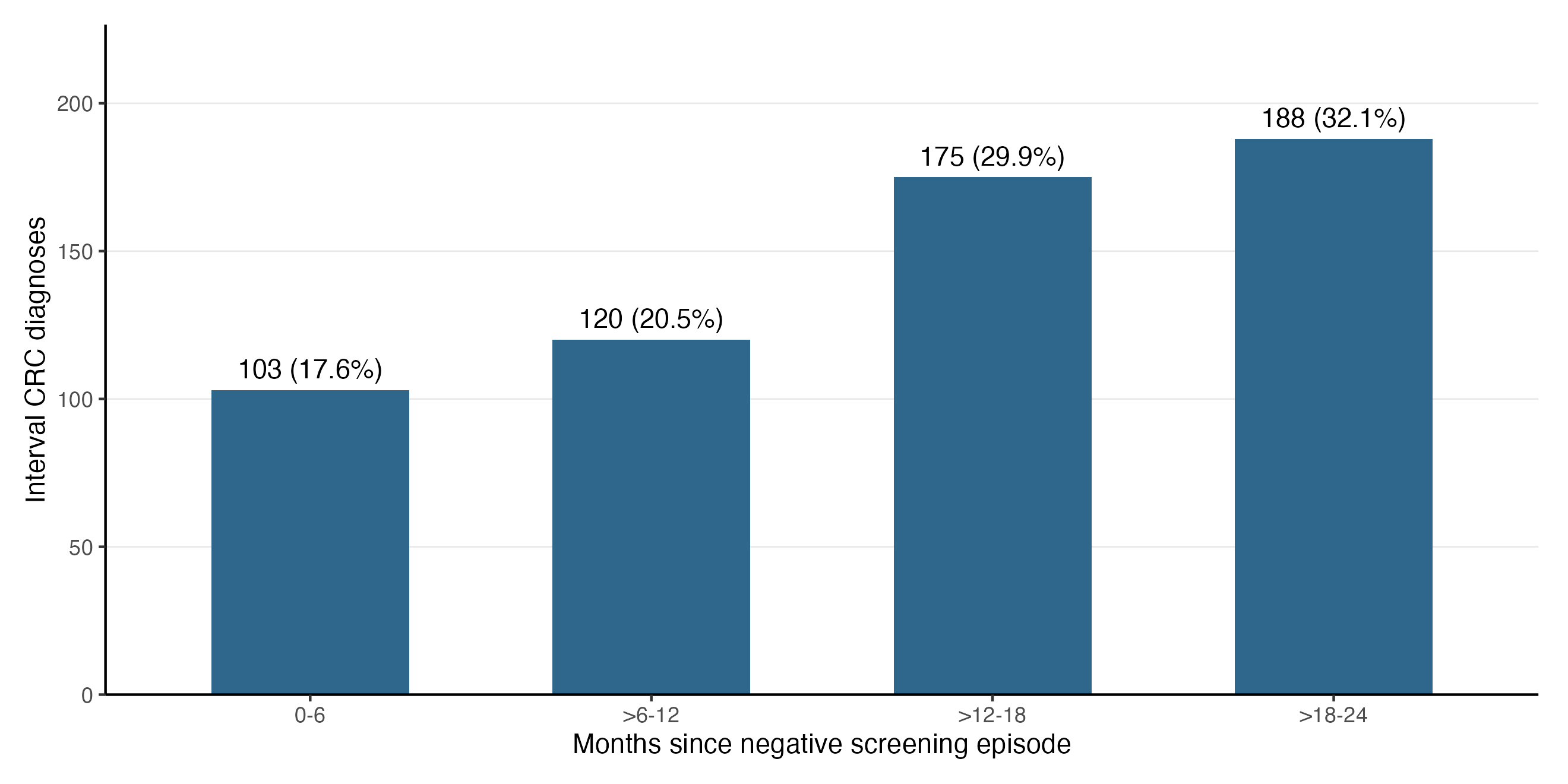

**Figure S4. Model-standardised cumulative incidence by f-Hb category**

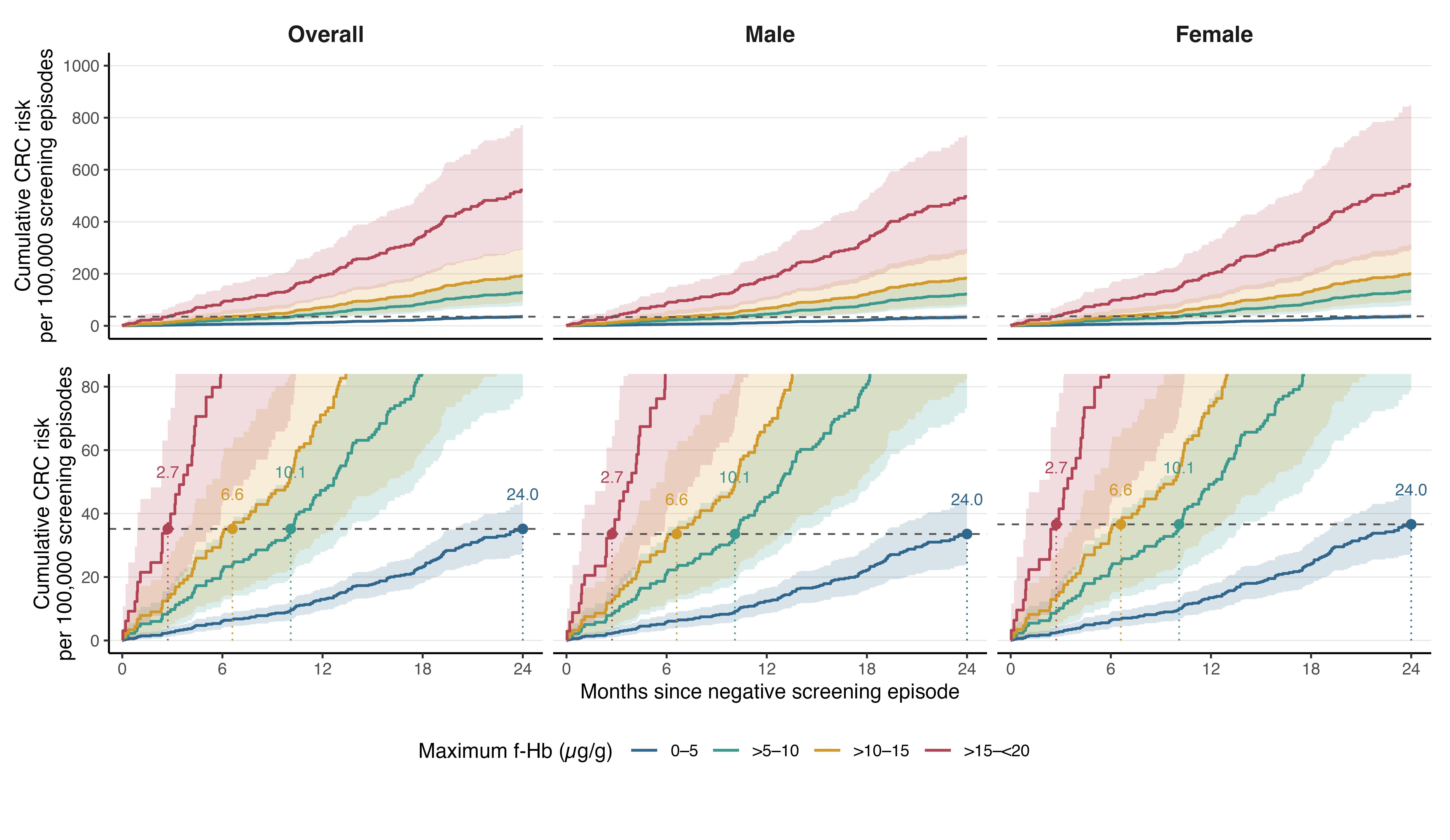

Risks per 100,000 screening episodes, standardised from paired cause-specific Cox models over the original overall or sex-specific covariate distribution. Shading shows pointwise 95% percentile confidence intervals from 2,000 participant-cluster bootstrap samples. The dashed line marks the panel’s 24-month risk at 0–5 µg/g; labels show first crossing times in months. Lower panels enlarge the reference-risk region.

**Figure S5. Distribution of negative-range f-Hb by five-year age group and sex**

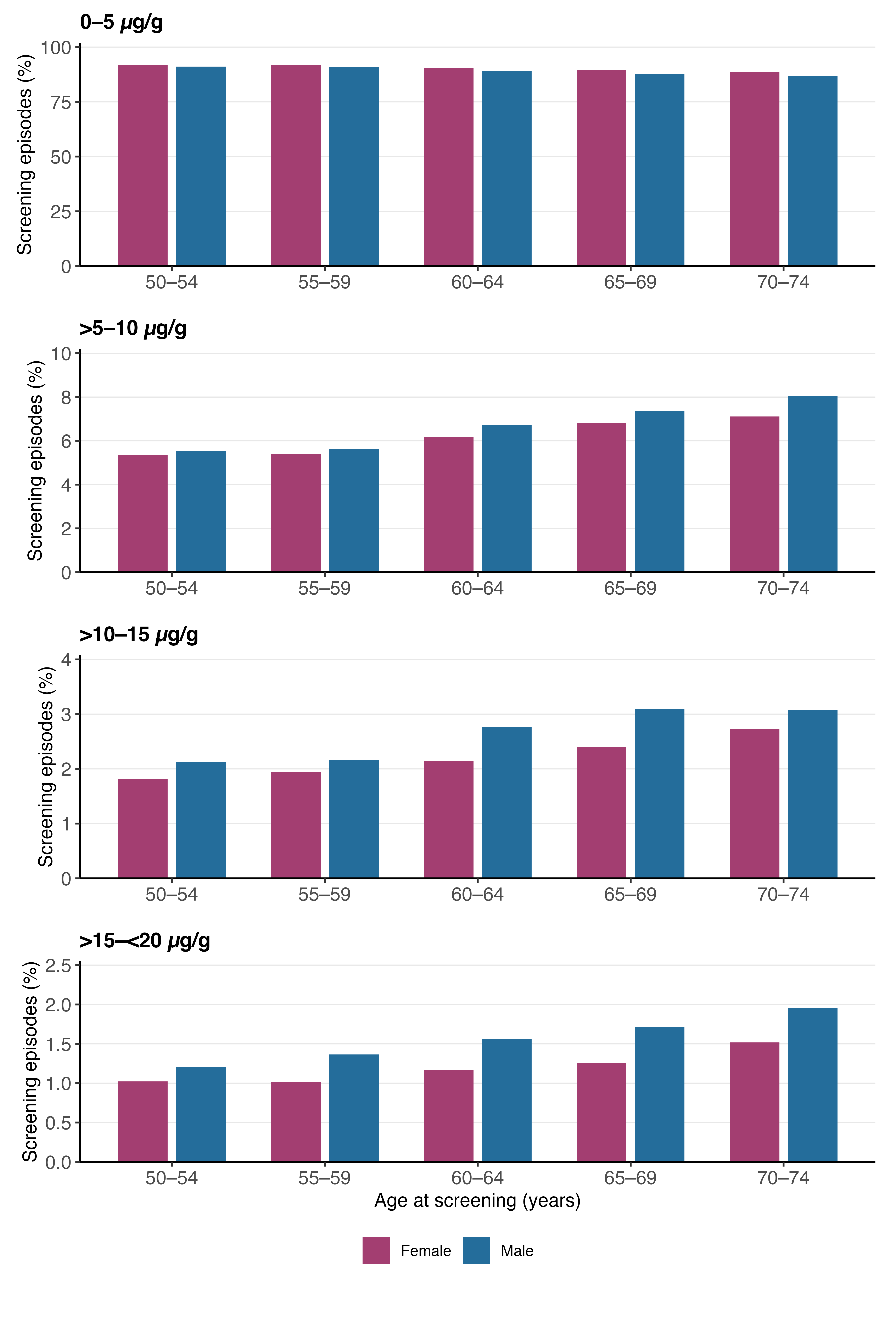
